# Discovery of Oncogenic Mediator Genes in Rectal Cancer Chemotherapy Response using Gene Expression Data from Matched Tumor and Patient-Derived Organoid

**DOI:** 10.1101/2024.01.29.24301906

**Authors:** Hanchen Huang, Chao Wu, Antonio Colaprico, Paulina Bleu, Wini Zambare, Janet Alvarez, Min Jung Kim, Aron Bercz, Lily Wang, Philip B. Paty, Paul B. Romesser, J. Joshua Smith, X. Steven Chen

**Author notes:** Equal contributions as co-first author (Hanchen Huang, Chao Wu). Co-corresponding authors Correspondence and requests for materials should be addressed to J.J.S., X.S.C.

## Abstract

Rectal cancer (RC) presents significant treatment challenges, particularly in the context of chemotherapy resistance. Addressing this, our study pioneers the use of matched RC tumor tissue and patient-derived organoid (PDO) models coupled with the innovative computational tool, Moonlight, to explore the gene expression landscape of RC tumors and their response to chemotherapy. We analyzed 18 tissue samples and 32 matched PDOs, ensuring a high-fidelity representation of the tumor bioloy. Our comprehensive integration strategy involved differential expression analyses (DEAs) and gene regulatory network (GRN) analyses, facilitating the identification of 5,199 genes governing at least one regulon. By using the biological processes (BPs) collected from Moonlight closely related to cancer, we pinpointed 2,118 regulator-regulon groups with potential roles in oncogenic processes. Further, through integration of Moonlight and DEA results identified 334 regulator-regulon groups significantly enriched in both tissue and PDO samples, classifying them as oncogenic mediators (OMs). Among these, four genes (NCKAP1L, LAX1, RAD51AP1, and NAT2) demonstrated an association with drug responsiveness and recurrence-free survival (RFS), offering new insights into the molecular mechanisms of chemotherapy response in RC. Our integrated approach not only underscores the translational fidelity of PDOs, but also harnesses the analytical prowess of Moonlight, setting a new benchmark for targeted therapy research in rectal cancer.

## Introduction

Colorectal cancer (CRC) is the second leading cause of cancer-related death in the US. In 2023, an estimated 153,020 US adults will be diagnosed with CRC[1]; RC makes up ∼1/3 of these cases, and incidence is rising in patients <50 years[2]. Due to tumor location and proximity to essential genitourinary organs, RC treatment is more challenging vs. colon cancer. Treatment for RC is trimodal, composed of 5-fluorouracil (5-FU)-based chemotherapy, neoadjuvant chemoradiation, and radical surgery (RS). As RS can cause functional deficits and impair quality of life (i.e., possible colostomy), modern trials have applied total neoadjuvant therapy (TNT) for locally advanced RC to optimize response – with TNT, all treatment is admistered upfront, before consideration of RS[3–5]. Some RC tumors respond completely to TNT (no residual tumor), and these patients can avoid RS (i.e., watch-and-wait approach)[3]. However, 50-60% of patients respond poorly to TNT (residual tumor) and require RS[6–8]. These patients also typically have worse survival compared to those with significant or complete resolution of tumor[4, 9]. With these significant issues, along with increasing incidence in young patients[10], there is a critical need to discover new therapeutic targets in TNT-resistant RC.

The efficacy of chemotherapy, which is the backbone of TNT, in treating cancer is significantly influenced by the complex interplay of various genetic factors, among which oncogenic mediator (MR) genes play a critical role. These genes, central to the pathways and networks involved in tumor progression, have profound implications for how cancer cells respond to chemotherapeutic agents. Oncogenic mediators often function within specific pathways, acting as crucial links in the transmission of oncogenic signals that contribute to drug resistance or sensitivity. Their ability to modulate key processes such as cell proliferation, apoptosis, DNA repair, and drug efflux mechanisms makes them pivotal in determining the success or failure of chemotherapy. Understanding the role of these mediators not only sheds light on the intricate molecular mechanisms underpinning chemotherapy resistance but also opens avenues for developing targeted therapies. By identifying and targeting these oncogenic mediators, it becomes possible to disrupt the cancer-specific pathways they govern, potentially enhancing the efficacy of chemotherapy and paving the way for personalized treatment strategies. This focus on oncogenic mediators thus represents a vital aspect of research in cancer therapeutics, where the ultimate goal is to tailor treatment approaches based on the unique genetic makeup of individual tumors, thereby maximizing therapeutic effectiveness while minimizing side effects.

Using the gene expression data from pateints or cell lines directly to discover MRs has limitations. For patients’ gene expression data, intratumoural heterogeneity (ITH) via stromal cells in the tumor microenvironment (TME) may hide subtle gene expression alterations associated with genetic diversity and heterogeneity within the tumor epithelium[11]. While cancer cell lines are a mainstay in preclinical research due to their ease of use and reproducibility, they have notable drawbacks, including the loss of tumor heterogeneity and genetic drift through long-time cell lien cultures, further diverging from the original tumor’s properties and potentially leading to less reliable or less clinically relevant results.There are also limited cell lines originating from rectal cancers, and verifying whether they truly come from the rectum or from patients who have received multimodal therapy is not feasible [12–14]. Finally, the complexity of genomic features (i.e., *curse of dimensionality* or *heterogeneity of input*) presents a key challenge for prediction and driver gene discovery tasks involving drug response[15–17]. This means the top genomic features selected from primary tumors may not be relevant in different patient cohorts partially due to the false positive signals from high-dimensional molecular data, which was also proved in CRC biomarker research[18–21].

To address the limitations of RC preclinical models, we built a biorepository from RC patient tumors and used them to generate the first viable *ex vivo* and *in vivo* RC tumoroid or patient-derived organoid (PDO) models[22] that retain histopathologic and clonal oncogenic mutations of the primary tumors. Importantly, these RC PDOs are predictive for patient TNT response, can be established from biopsy material (90% success rate), and can be derived *both pre-treatment and after TNT* in RC tumors that fail to respond and require radical surgery[22].

Recently, we developed an innovative computational biology tool, Moonlight, to discover cancer driver genes, including oncogenes, tumor-suppressor genes[23]. Moonlight relies on observations of experimentally validated cancer driver genes whose expression is modulated in cellular assays, together with the quantification of process markers, such as cellular proliferation, apoptosis, and invasion. To accomplish this task, we curated >100 biological processes linked to cancer, including proliferation and apoptosis. During this manual curation, we provided Moonlight information on whether the activation of each process leads to promotion or reduction of cancer. Once Moonlight identifies an oncogenic process altered in tumors using gene expression data, it detects genes that activate or inhibit this process, which are defined as oncogenic mediators. This method has been successfully applied to >8,000 tumor samples from 18 cancer types to discover cancer driver genes[23]. We also discovered inactivation of tumor suppressors and that tissue type and subtype could indicate dual role status. These findings help explain tumor heterogeneity and could guide therapeutic decisions[23].

We conducted the current study by integrating RC PDO, tumor tissue, and drug response data using Moonlight to identify novel MR gene targets with the goal of rendering resistant tumors sensitive to TNT. The PDOs, derived from rectal cancer tumors, not only preserve the histopathological and genetic characteristics of the primary tumors, but also demonstrate predictive value for patient response to TNT. These organoid models offer a more accurate and dynamic representation of the tumor microenvironment, overcoming the challenges of intratumoral heterogeneity and the limitations of cell line models. Coupled with our innovative computational tool, Moonlight, which effectively identifies oncogenic mediators by analyzing regulatory networks and biological processes implicated in cancer, our approach provides a robust platform for uncovering novel therapeutic targets. By integrating transcriptomic data from RC PDOs, tumor tissue, and drug response, this study seeks to identity and characterize oncogenic mediators, enhance TNT efficacy and improve survival outcomes for rectal cancer patients.

## Results

### Integration of gene expression data from matched RC tumors and PDOs

In this study, we employed a multi-tiered, proof-of-principle integration strategy to unravel the complex molecular landscape of rectal cancer and its response to chemotherapy as illustrated in **Fig 1**. Our rationale is that 5-fluorauracil-based chemotherapy has been and remains the backbone of chemotherapy and chemoradiation in rectal cancer for the past 60 years – however, no new therapies have been added that specifically address the patients with chemoresistant tumors[3, 24–27]. Our approach began with a comprehensive analysis of transcriptome data from 18 RC tissue samples and matched patient-derived organoids (PDOs), focusing on the identification of drug-related genes based on IC50 outcomes according to FOLFOX (5-fluorouracil, leucovorin, and oxaliplatin) treatment of RC PDOs from our RC biorepository. IC50 in RC PDOs has been shown to correlate well with both oncologic and clinical outcomes [22, 28]. The integration extended to a thorough examination of The Cancer Genome Atlas (TCGA) COAD/READ data, where differential expression analyses (DEAs) were conducted to distinguish between tumor and paired normal tissues, uncovering key differentially expressed genes (DEGs). A subsequent gene regulatory network (GRN) analysis provided a structural and functional framework for understanding the interactions and regulatory relationships among these genes. Central to our strategy was the application of Moonlight, a tool adept at analyzing over a hundred biologically relevant processes, to identify altered oncogenic processes in tumors. This analysis pinpointed specific oncogenic mediator genes acting as regulatory hubs within these processes. By amalgamating the insights gleaned from RC tumors and PDOs, TCGA data, and the process-specific gene evaluation via Moonlight, we established a robust, multidimensional approach. This approach not only enhanced our understanding of the oncogenic pathways in rectal cancer but also illuminated potential oncogenic mediators, offering putative targets for sensitizing resistant tumors to chemotherapy.

**Figure 1.**
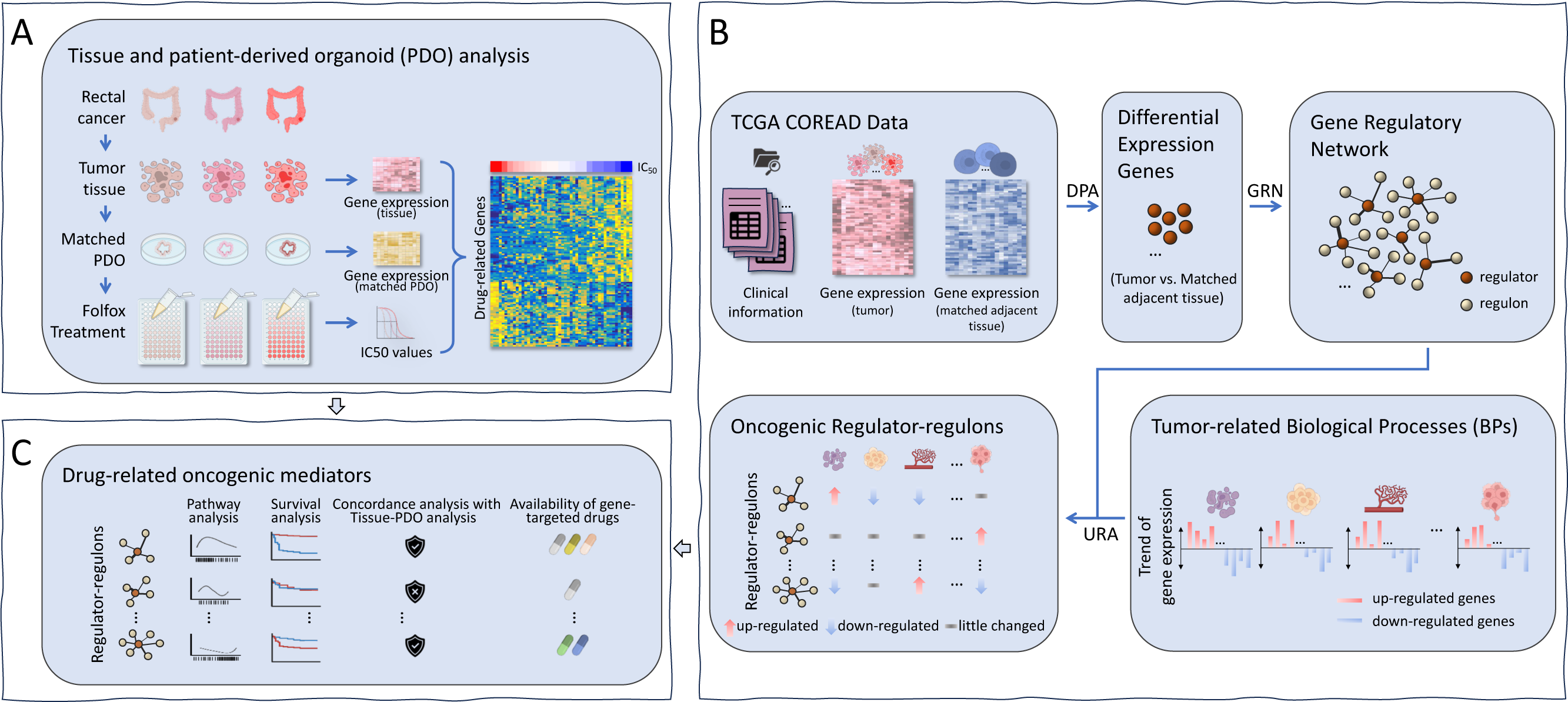
The workflow of the study. It consists of (A) tissue-organoid analysis for screening genes associated to drug-response, (B) Moonlight-based analysis for discovering oncogenic regulator-regulons groups of colorectal cancer, and (C) the integrated analysis for identifying oncogenic mediators as potential indicators of drug responsiveness.

### Gene expression consistency between cancer tissues and matched PDOs

In our analysis, all 50 samples, comprising 18 tissue samples and 32 organoid samples, met the stringent criteria set for quality control and were subsequently included in further analysis (put these criteria in Supplementary Material?). Following the removal of genes with minimal expression (detailed in the Methods section), a substantial pool of 17,418 genes was retained for analysis. Initial evaluations focused on the consistency of gene expression within organoid samples derived from individual patients (**Fig. 2A**). For each set of three replicated organoid samples per patient, a remarkable consistency was observed, with pairwise Spearman’s correlation coefficients invariably exceeding 0.958 (P-value < 0.001), underscoring the reliability of organoid models in reflecting patient-specific gene expression profiles.

**Figure 2.**
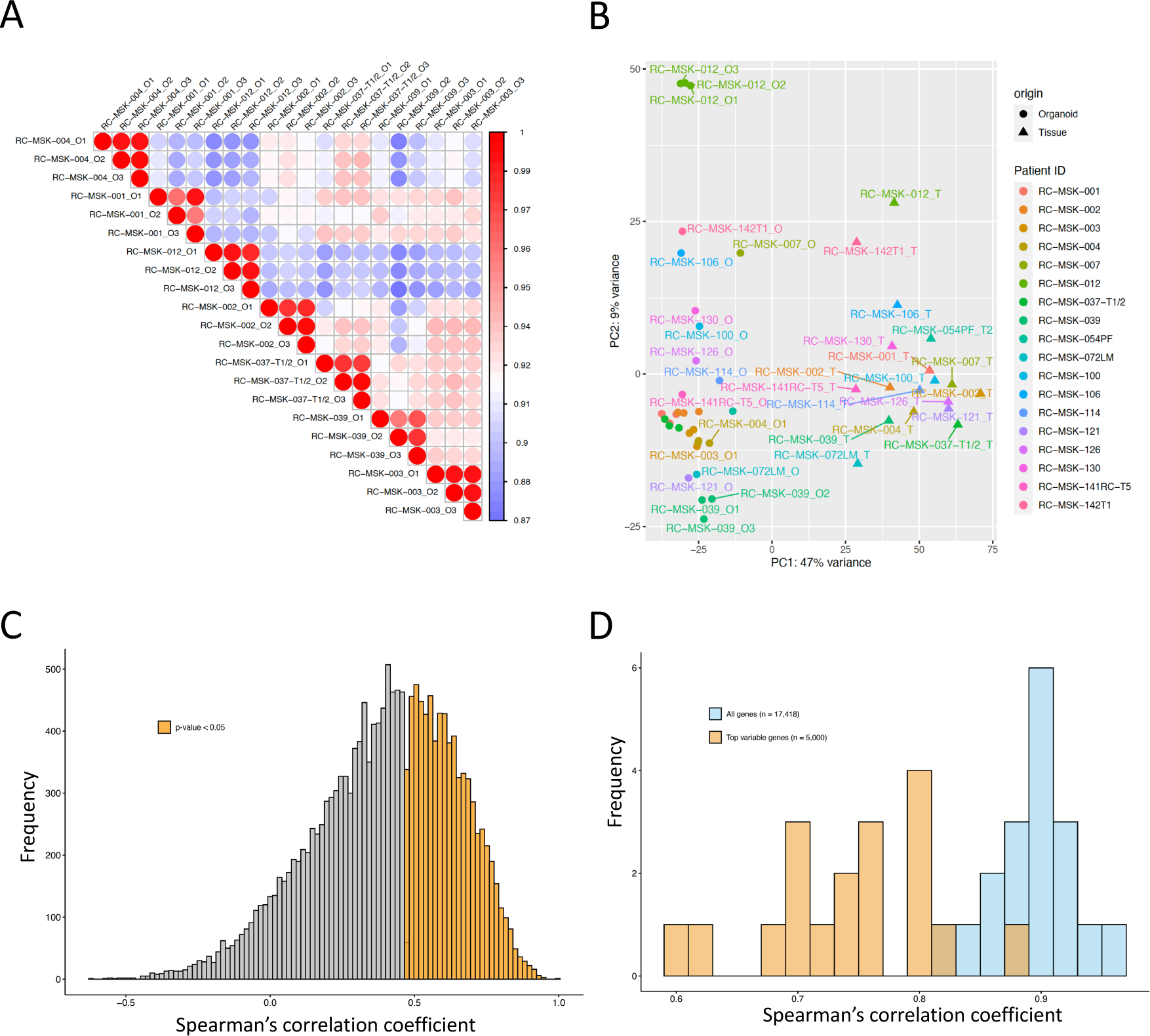
Reproducibility of patient-derived organoid and tissue-organoid correlation. (A) Sample-wise correlation based on 21 organoid samples derived from seven patients. (B) Principal component analysis (PCA) of 18 tissue and 32 matched organoid samples based on gene expression data. (C) Histograms of gene-wise based on expression data of all genes, and (D) sample-wise correlation coefficients based on all genes (blue) and top-5000 variable genes selected from organoid (orange) of 18 tissue-organoid matched patients.

Further, Principal Component Analysis (PCA) was employed on the gene expression data from the 50 samples, revealing a predominant segregation into two distinct clusters, one encompassing tissue samples and the other comprising organoid samples (**Fig. 2B**). Notably, the organoid cluster exhibited reduced variability along the first principal component (PC1), but heightened variability along the second principal component (PC2), suggesting a nuanced representation of genetic diversity within these models. Moreover, replicated organoid samples from the same patients were observed to cluster tightly on the PCA plot, reinforcing the reproducibility of the organoid model.

We then probed the correlation in gene expression between primary tumor tissues and their derivative organoid samples (**Fig. 2C**). In scenarios involving multiple organoid samples from a single patient, the gene expression for the organoids was represented by the arithmetic mean of the samples. A substantial proportion of genes, 16,918 out of 17,418 (97.1%), demonstrated a positive correlation (Spearman’s rho > 0) between tissue and organoid gene expressions. Within this subset, 6,514 genes manifested a statistically significant correlation, marked by a Spearman’s rho > 0.5 and a P-value < 0.05 (**Supplementary Table 1**). A patient-specific analysis was further conducted, assessing the gene expression correlation between tissue samples and their corresponding organoid samples, considering the full spectrum of genes as well as a subset of highly variable genes (**Fig. 2D**). This analysis consistently revealed high correlation coefficients across all patient samples, affirming the organoid model’s capacity to faithfully recapitulate the gene expression landscape of the original tumor tissues.

### Gene selection for drug responsiveness

To discern genes correlated with drug response, we executed regression analyses on tumor tissues and their corresponding organoids, paired with IC50 values from FOLFOX treatment (see Methods section). This analysis identified a total of 660 genes from tissues (refer to **Supplementary Table 2**) and 819 genes from organoids (refer to **Supplementary Table 3**), each exhibiting a significant correlation with drug response (p-value < 0.05). Notably, there was an overlap of 75 genes between the two datasets (**Fig. 3A** and itemized in **Supplementary Table 4**). A subsequent sample-wise correlation assessment demonstrated a high level of consistency in the expression of these drug-related genes between tissue and organoid samples, as depicted in **Fig. 3B**. Of the 75 drug response genes (DRGs) highlighted, 17 were positively linked to drug resistance, while 58 were positively linked to drug sensitivity, as shown in **Fig. 3C**. Utilizing the expression profiles of these DRGs, unsupervised hierarchical clustering was performed on a combined dataset of tissue and organoid samples. This clustering effectively segregated the samples into two distinct groups, each exhibiting contrasting patterns of drug responsiveness. Furthermore, recognizing the limited sample size in this analysis and aiming for broader applicability in integrated analyses, we also compiled a supplementary list of 458 genes. These genes exhibit a marginally significant correlation with drug response in both tissue and organoid samples, as determined using a relaxed threshold of p-value 0.15 (see **Supplementary Table 5**).

**Figure 3.**
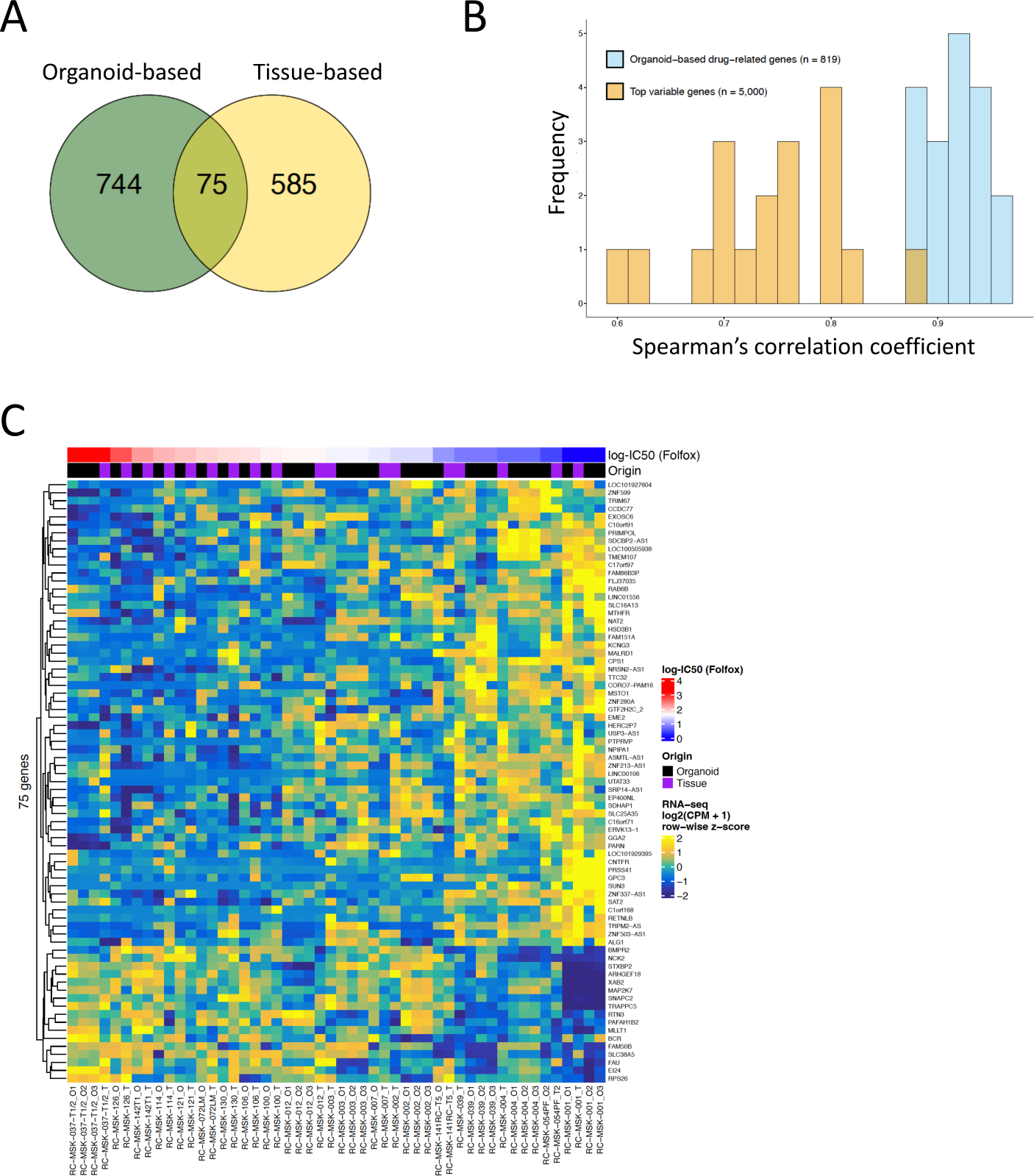
Identification of drug responsiveness genes. (A) Venn’s diagram of drug-related genes (P < 0.05) identified from organoid (green, *n* = 819) and tissue (yellow, *n* = 660) samples. (B) Histograms of sample-wise correlation coefficients based on top-5000 variable genes (orange) and 819 drug-related genes (blue) identified from organoid samples. (C) Heat map of gene expression of 75 drug-related genes (P < 0.05 in both tissue and organoid samples), sorted by descending order based on log-IC50 values of Folfox treatment in sample-wise.

### Detection of oncogenic regulator-regulons through Moonlight

In our study, we leveraged an enhanced Moonlight workflow to systematically screen for potential oncogenic mediators from the combined TCGA dataset of rectal (READ) and colon (COAD) cancers, acknowledging the relatively limited sample size in each category (**Fig. 4A**). A cohort of 223 patients, each with matched primary tumor and adjacent normal tissue samples, and documented chemotherapy history, formed the basis of our analysis. Post exclusion of microRNA data, the remaining 17,082 protein-coding genes underwent a Differential Phenotype Analysis (DPA) using Moonlight, which yielded 5,272 genes demonstrating significant differential expression (log2|FC| > 1 and FDR < 0.01, **Supplementary Table 6**). Subsequent Gene Regulatory Network (GRN) analysis within Moonlight scrutinized these differentially expressed genes (DEGs) as potential regulators, exploring their associated regulons from the entire gene pool of 17,082 candidates. This comprehensive analysis identified 5,199 genes that govern at least one regulon (**Supplementary Table 7**), with the most influential gene regulating up to 783 regulons.

**Figure 4.**
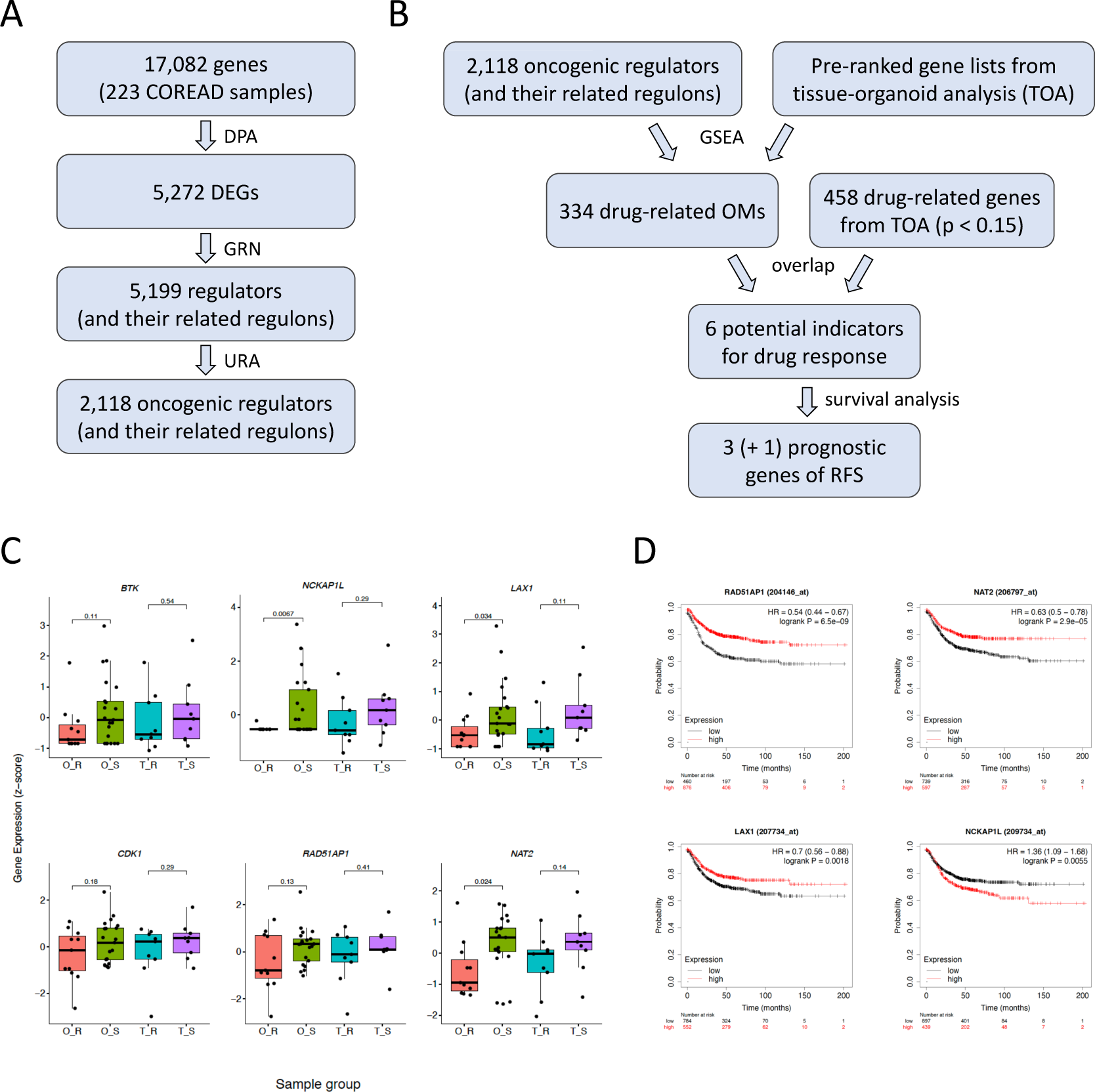
Identification of potentially oncogenic mediators via (A) Moonlight and (B) integrated analysis. (C) Boxplots of six genes as potential indicators for drug response, and (D) four genes out of the six showing significant association with recurrence-free survival (RFS) among colon cancer patients (*n* = 1,296) from multiple datasets.

To delve deeper into the biological significance of these findings, we conducted modified functional enrichment analysis (FEA) and upstream regulator analysis (URA) using the unique modules from Moonlight, pinpointing regulator-regulon groups implicated in cancer-relevant biological processes (BPs). Initially, Moonlight offered insights into 101 BPs; however, we honed our focus to 25 BPs intimately associated with cancer progression, such as “apoptosis,” “tumor cell line proliferation,” and “tumor growth” (**Supplementary Table 8**). These BPs were dissected further using Ingenuity Pathway Analysis (IPA) to isolate a subset of genes with strong associations to each process. An automated PubMed literature review annotated these genes, hinting at the direction of their expression alterations during these processes. In this refined FEA and URA, our criteria for significance were stringent: we considered only regulator-regulon groups with a Moonlight |Z-score| over 1 and a Fisher’s exact test P-value below 0.05. This approach led us to identify 2,118 regulator-regulon groups, each potentially playing a pivotal role in cancer-related BPs (**Supplementary Table 9**), marking a crucial step in understanding the regulatory underpinnings of oncogenic processes in rectal and colon cancers.

### Integration of the discoveries from Moonglight and tissue-organoid analyses

In our pursuit to discern specific associations between the 2,118 cancer-related regulator-regulon groups and drug responsiveness, we embarked on an integrated analysis, as depicted in **Fig. 4B**. This process commenced with a Gene Set Enrichment Analysis (GSEA), wherein the regulator-regulon groups were examined alongside the insights derived from our tissue-organoid investigation. Employing the 2,118 regulator-regulon groups as gene sets, the GSEA utilized two pre-ranked gene lists, each organized in descending order based on the regression’s t-value, one for tissue and the other for organoid analyses. To enhance the robustness of our findings, we focused on 11,198 genes that consistently indicated drug responsiveness trends across both tissue and organoid samples. This GSEA identified 334 regulator-regulon groups that exhibited significant enrichment across both modalities, leading us to classify these 334 entities as oncogenic mediators (OMs). In our analysis, six genes – BTK, NCKAP1L, LAX1, CDK1, RAD51AP1, and NAT2 – emerged as noteworthy for their association with drug responsiveness. Although the threshold for significance was set at a more inclusive level (P-value < 0.15), these genes consistently showed a positive correlation with sensitivity to FOLFOX treatment in both tissue and organoid samples, as demonstrated in **Fig. 4C**. This suggests a potential link between these genes and chemotherapy response.

To further determine if these genes were associated with an oncologic outcome, we tested the prognostic potential of these six OMs using the Kaplan-Meier Plotter database, with results illustrated in **Fig. 4D**. This analysis revealed that RAD51AP1, NAT2, and LAX1 were significantly associated with extended recurrence-free survival (RFS), showcasing HR values of 0.54 (logrank P = 6.5e-09), 0.63 (logrank P = 2.9e-05), and 0.70 (logrank P = 1.8e-03), respectively. These findings align well with the drug responsiveness trends observed in our integrated analysis. Interestingly, NCKAP1L, despite showing a significant association with RFS (HR = 1.36, logrank P = 5.5e-05), exhibited a trend that diverged from the drug response analysis.

To delve into the therapeutic prospects of the identified drug-related genes, we assessed the possibility of targeting specific perturbagens against these genes through the Connectivity Map tool. This exploration led to the identification of 89 perturbagens targeting 24 out of the 334 oncogenic mediators and 107 drugs targeting 23 out of the 458 drug-related genes (p < 0.15), as highlighted in our previous tissue-organoid analysis (**Fig. 5A and 5B; Supplementary Table 11 and 12**). This revelation underscores the potential for therapeutic intervention and paves the way for further investigation into the efficacy of these perturbagens in modulating drug responsiveness in rectal cancer.

**Figure 5.**
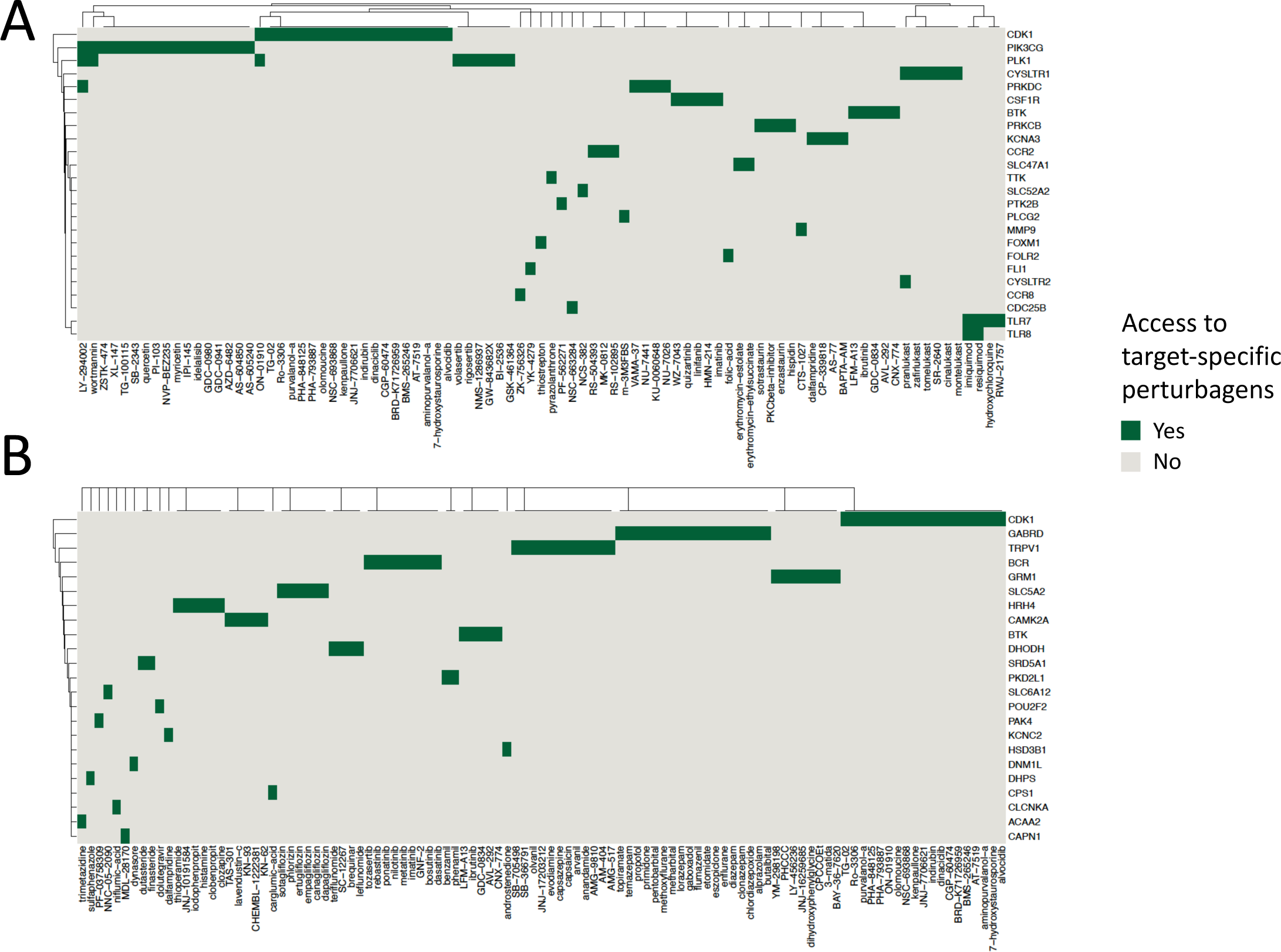
Heatmap showing availability of perturbagens from the Connectivity Map in columns and their target drug-related genes in rows from (A) Moonlight-based oncogenic mediators and (B) tissue-organoid analysis with P < 0.15.

## Methods

### Human RC tissue and patient-derived organoid

A total of 18 patients diagnosed with rectal cancer (RC) at Memorial Sloan Kettering Cancer Center were selected in our study. For each patient, one tumor tissue was obtained before treatment, and at least one matched patient-derived organoid (PDO) samples were generated. In specific, for each of the eleven out of 18 patients, one single PDO was generated, while to validate the replicability of PDO model, each of the remaining seven patients had triplicated organoid samples. Endoscopic tumor biopsies obtained with biopsy forceps were obtained from RC patients undergoing standard of care diagnostic biopsy or on-treatment biopsy in the outpatient setting. The specimens are collected per protocol and tissue was prioritized as follows: sample sent to pathology, sample for PDO, and remaining saved for banking. Tissues are chopped into ∼1-mm pieces in cold PBS + antibiotics, and after tissue processing, tumor cells are resuspended in 300 μL of Matrigel and plated onto 24-well culture plates (40 μL droplet per well). After the gel solidifies, 700-800 μL of culture medium is added to wells and incubated at 37°C. PDOs are established and maintained in Matrigel as described in Ganesh et al.[22]. Consequently, 18 tissues and 32 matched organoids were involved in this study and were then RNA-sequenced (see below).

### Preparation and RNA-sequencing of tissue and organoid samples

Surgically resected rectal tissue or biopsy tissue was washed with ice-cold PBS-Abs buffer (phosphate-buffered saline with antibiotic-antimycotic (Gibco™), gentamicin (Gibco™) and plasmocin (Invivogen). The tissue was divided into two parts. One part was flash-frozen in liquid nitrogen and then reserved at -80 °C for later purposes. The other part was processed for tumoroid culturing purposes, as reported[22]. The patient-derived tumoroids were cultured and expanded in Matrigel for three passages. To RNA, well-growing tumoroids in 4-6 matrigel domes (40ul matrigel/dome) were dissociated with TrypLE Express (Gibco™) as reported[22]. The obtained pellet was further washed with 5 ml of ice-cold PBS. Then, RNA was extracted from the tumoroid pellet using the AllPrep RNA kit (Qiagen) following the manufacturer’s instructions. If the RNA needs to be extracted separately, then the PureLink™ RNA Mini Kit (Invitrogen™) was used. The tissue RNA was extracted from the flash-frozen specimen using the AllPrep RNA kit.

For RNA sequencing, total RNA was prepped with the Nugen Universal Plus mRNA-Seq (M01442 v2) using 750ng via Qubit and 14 cycles PCR. Libraries were sequenced on Illumina Novaseq 6000 and ∼50M PE150 were generated per sample.

### Tissue-organoid data analysis

To process raw FASTQ-formatted RNA sequencing data from organoid and tissue samples, we utilized the nf-core/rnaseq tool[29] (version 3.12.0) in Nextflow pipeline[30] (version 22.10.7) with default settings and parameters. The data processing workflow consisted of pre-processing quality control (QC), STAR-based reads alignment to the reference genome (Genome assembly GRCh38: https://www.ncbi.nlm.nih.gov/datasets/genome/GCF_000001405.26/), and Salmon-based gene expression data quantification. Detailed information of RNA-seq analysis pipeline of Nextflow could be referred to at https://nf-co.re/rnaseq/3.12.0.

Data analysis for count-based gene expression data were then performed using R software[31] (version 4.2.2) in RStudio[32] (version 2023.09.0). We transferred count data to count-per-million (CPM) using edgeR package[33] (version 3.40.2), and retained only unduplicated genes with CPM values simultaneously greater than 0.1 among at least 15% tissue samples, 15% organoid samples and 15% tissue plus organoid samples. We implemented limma-based standardization using calcNormFactors function from edgeR package, and applied a log2 transformation to the CPM values, resulting in normalized data for the downstream analysis.

To validate the reproducibility of the triplicated organoid samples, we created heatmaps to visually represent the Spearman’s correlation matrix. To evaluate the correlation between gene expression in organoid and tissue samples, we computed patient-wise Spearman’s correlation coefficients. For patients with replicated organoid samples, we averaged their gene expression values to align with tissue values. Histograms were then generated to illustrate the distribution of Spearman’s correlation coefficients for all genes and top-5000 variable genes.

To identify genes associated with drug responsiveness (Fig. 1A), we conducted a preliminary screening through differential expression analysis (DEA) using regression models. As multiple replicated samples were generated for a proportion of organoids, we treated the patient identifier as a mixed-effect variable for regression modeling. Specifically, we fitted the model *log_2_(CPM + 1) ∼ 1|patient.id + ln(IC_50_)* using the lmer function in lme4 R package for organoid samples. For tissue samples, the model *log_2_(CPM + 1) ∼ ln(IC_50_)* was fitted using the limma-voom method from the limma package [34]. Here, gene expression was measured by log2-transformed CPM. Genes with unadjusted P-values < 0.05 were selected as candidates associated with chemotherapy response.

Furthermore, we identified the intersection of candidate genes from both organoid and tissue data and used them to generate a heatmap utilizing the ComplexHeatmap[35] package.

### A modified Moonlight workflow

To identify a set of oncogenic genes based on TCGA cohort[36], we performed customized analysis based on Moonglight pipeline[37] (Fig. 1B). For all human colon (COAD) and rectal (READ) cancer patients from TCGA, high-throughput RNA-seq data normalized by upper quartile (HTseq – FPKM-UQ) and clinical information were downloaded and processed with TCGABiolinks package[38]. To prioritize the investigation of drug response, only patients who received chemotherapy were included in the analysis. For the patients with duplicated primary solid tumor (TP) samples, only the first sample profiled by date was retained. For consistency with our dataset, we excluded genes that encode microRNAs from subsequent analysis.

The MoonlightR::DPA module was employed to identify differentially expressed genes (DEGs) between tumor and normal tissues, with a significance threshold set at false discovery rate (FDR) < 0.01 and log_2_|FC| > 1. To uncover potential target regulons associated with each DEG, the MoonlightR::GRN module was utilized to construct a gene regulatory network (GRN) based on mutual information. This GRN comprised regulator-regulon groups, where each DEG was considered a potential regulator, and its regulons were selected from the entire gene pool using algorithms in the GRN module. To further investigate genes associated with cancer-related biological processes (BPs), a subset of BPs was chosen from the original 101-BP list predefined in the Moonlight R package. Within each BP, there was a set of genes that could potentially exhibit upregulation, downregulation, or unspecified changes in expression levels during that specific BP. A selective subset of cancer-related BPs was retained for subsequent analysis in MoonlightR::URA. In this step, the enrichment significance of each combination of regulator-regulon group and BP was assessed using URA-derived Moonlight Z-scores and P-values. Regulator-regulon groups with at least one significantly enriched BP, meeting the threshold of Mooonlight |Z| > 1 and P < 0.05 in the URA step, were selected as candidates for further analysis.

Gene set enrichment analysis (GSEA) using the fgsea[39] package was performed for each regulator-regulons group to integrate the Moonlight-based analysis and tissue-organoid analysis (Fig. 1C). A set of regulons, representing the potentially target genes for each regulator, was utilized as a gene set. Two gene lists, one from organoid analysis and the other from tissue analysis, were employed in GSEA. These lists were ranked in descending order of log_2_FC. Only genes exhibiting the same trends in drug response were retained for GSEA in both pre-ranked lists. The gene set size was constrained, with a maximum of 500 genes and a minimum of 5 genes.

Regulators from the regulator-regulons groups that demonstrated significant enrichment patterns (P-value < 0.05) in both organoid-based and tissue-based lists were ultimately selected as oncogenic mediators (OMs) through this integrated analysis.

### Survival analysis

To investigate the prognostic performance of genes, we retrieved an online resource namely Kaplan-Meier Plotter (https://kmplot.com/analysis/index.php?p=service&cancer=colon), which provided survival analysis for 1,296 patients with colon cancer from 16 datasets based on their gene expression and survival information [40]. For each gene, patients were divided into two groups, using an automatic classification with the optimal cutoff in gene expression. This method aimed to achieve the lowest log-rank p-value for recurrence-free survival (RFS) between the groups, and based on which, a Kaplan-Meier (KM) plot was generated for each gene. When multiple probes corresponded to the same gene, the probe with the smallest p-value was selected as the representative.

### Connectivity Map analysis

We used Connectivity Map[41] online tools to retrieve potential compounds and perturbations for genes of interest. A text file namely compoundinfo_beta.txt containing a full list of perturbagens and their target genes was downloaded via https://clue.io/data/CMap2020#LINCS2020.

## Discussion

In this study, the innovative integration system we developed and employed represents a significant leap forward in the field of rectal cancer (RC) research and current treatment modalities, where the backbone of treatment has remained the same (even for resistant tumors) for more than 70 years. Our data is particularly important relative to better understanding and predicting chemotherapy response. This system highlights the strengths of using matched tumor tissue and PDO models with advanced computational analysis, specifically through the Moonlight tool to uncover pivotal oncogenic mediators (OMs) influencing chemotherapy response.

The remarkable alignment in gene expression patterns between RC tumors and their derived PDOs, as evidenced in our study, underscores the robustness and translational fidelity of the PDO model. This fidelity is not merely a quantitative match of gene expression levels, but reflects a deeper, more nuanced replication of the tumor microenvironment and its inherent genetic complexities. The substantial positive correlation in gene expression between primary tumor tissues and their corresponding organoid samples, as indicated by 97.1% of genes showing a positive Spearman’s rho (16,918 out of 17,418 genes), reinforces the reliability of the organoid model in reflecting the genetic landscape of individual tumors. Notably, a significant number of these genes (6,514) not only showed positive correlation but also reached a level of statistical significance (Spearman’s rho > 0.5, P-value < 0.05). This underscores the organoids’ ability to accurately replicate the gene expression patterns of the original tumor tissues, as further evidenced by the consistent high correlation across all patient samples in our analysis.

This congruence is of paramount importance, particularly considering the challenges posed by intratumoral heterogeneity and the dynamic nature of tumor evolution. Traditional in vitro models and cell lines have often fallen short in capturing this heterogeneity, leading to gaps in translation from bench to bedside. However, the matched gene expression data from RC tumors and PDOs in our study bridge this gap, offering a more accurate and clinically relevant model for studying tumor biology and drug response.

Moreover, the consistency in gene expression profiles reinforces the reliability of PDOs not only in reflecting the genetic makeup of the primary tumor, but also in their potential to predict therapeutic responses. This predictive capacity is crucial for precision medicine, where understanding the individual behavior of each tumor can guide tailored treatment strategies, potentially improving patient outcomes.

Furthermore, the matched gene expression data provide an invaluable resource for exploring the molecular underpinnings of RC. They enable a detailed dissection of oncogenic pathways, identification of potential biomarkers, and elucidation of drug resistance mechanisms. This depth of understanding is instrumental in advancing the development of targeted therapies and improving the efficacy of existing treatment modalities.

In our study, the utilization of Moonlight as an analytical tool has significantly enhanced our understanding of the complex gene regulatory networks involved in rectal cancer. The ability of Moonlight to dissect extensive gene expression data and pinpoint pivotal oncogenic mediators (OMs) has been instrumental in unveiling the intricate molecular interactions that govern tumor behavior and chemotherapy response. This computational tool goes beyond traditional differential expression analysis, offering a nuanced perspective by identifying regulator-regulon groups and mapping out the landscape of biological processes implicated in cancer progression. The insights gained from Moonlight analysis are not merely of academic interest; they hold profound implications for the development of targeted therapies. By spotlighting the key OMs within the regulatory networks, Moonlight provides actionable targets for therapeutic intervention, potentially ushering in a new era of precision medicine in rectal cancer treatment where strategies are tailored based on the unique molecular signature of each tumor.

The identified four OMs, RAD51AP1, NAT2, and LAX1, have demonstrated a marked association with extended recurrence-free survival (RFS), suggesting their potential as prognostic markers for favorable treatment outcomes. Notably, RAD51AP1, known for its role in DNA repair and maintenance of genomic stability, underscores the critical interplay between DNA damage response mechanisms and chemotherapy efficacy. The correlation of RAD51AP1 with improved RFS aligns with its function in facilitating accurate DNA repair, a process that, when compromised, can lead to therapy resistance.

Similarly, NAT2, with its involvement in drug metabolism, highlights the importance of individual genetic variations in modulating the pharmacokinetics and pharmacodynamics of chemotherapeutic agents. The association of NAT2 with RFS reflects the potential impact of metabolic pathways on drug response, pointing to the need for personalized therapeutic strategies that consider metabolic profiling.

The association of LAX1 with chemotherapy response and RFS introduces intriguing questions about its role in cancer biology. While less is known about LAX1 compared to RAD51AP1 and NAT2, its correlation with treatment outcomes invites further investigation into its biological function and potential as a therapeutic target.

Interestingly, NCKAP1L demonstrated a significant association with RFS, but displayed a trend opposite to that observed in the drug response analysis. This paradoxical finding indicates the complex nature of cancer biology and the multifaceted roles that genes can play in different contexts of tumor development and treatment response.

These findings emphasize the intricate relationship between oncogenic mediators and chemotherapy response in RC. The identified OMs not only offer insights into the molecular underpinnings of drug sensitivity and resistance, but also hold promise for the development of novel therapeutic targets and treatment strategies. By focusing on the unique genetic makeup of individual tumors and considering the functional roles of these OMs, we can pave the way for more personalized and effective treatment approaches, ultimately improving patient outcomes in rectal cancer.

## Data Availability

All data produced in the present study are available upon reasonable request to the authors

## List of Supplementary Tables

Supplementary Table 1 | Genes with significantly positive correlation between tissues and organoids.

Supplementary Table 2 | Genes with significant association with drug response in tissue samples (P < 0.05).

Supplementary Table 3 | Genes with significant association with drug response in organoid samples (P < 0.05).

Supplementary Table 4 | Genes with significant association with drug response in both organoid and tissue samples (P < 0.05).

Supplementary Table 5 | Genes with marginally significant association with drug response in both organoid and tissue samples (P < 0.15).

Supplementary Table 6 | Differentially expressed genes (DEGs) between tumor and normal tissues in TCGA-COREAD cohort (P < 0.05).

Supplementary Table 7 | Gene Regulatory Network (GRN) built by Moonlight based on TCGA-COREAD cohort.

Supplementary Table 8 | Biological Processes (BPs) that are selected for upstream regulator analysis (URA).

Supplementary Table 9 | Regulator-BP pairs from Upstream Regulator Analysis (URA) with |Moonlight’s Z| > 1 and P < 0.05.

Supplementary Table 10 | Oncogenic Mediators (OMs) identified by GSEA in the integrated analysis (P < 0.05).

Supplementary Table 11 | Available perturbagens (compounds) targeting drug-related OMs in Connectivity Map (CMAP) project.

Supplementary Table 12 | Available perturbagens (compounds) in CMAP targeting genes that are marginally associated with drug response (P < 0.15) in both organoid and tissue samples.

## DECLARATIONS

### Availability of data and materials

The datasets analyzed in this study are avaialbe upon the request to the corresponding authors

